# HIGH VERSUS STANDARD DOSES OF CORTICOSTEROIDS IN COVID-19 PATIENTS WITH AN ACUTE RESPIRATORY DISTRESS SYNDROME: a controlled observational comparative study

**DOI:** 10.1101/2020.07.17.20156315

**Authors:** Enric Monreal, Susana Sainz de la Maza, Elena Natera-Villalba, Álvaro Beltrán-Corbellini, Fernando Rodríguez-Jorge, Jose Ignacio Fernández-Velasco, Paulette Walo-Delgado, Alfonso Muriel, Javier Zamora, Araceli Alonso-Canovas, Jesús Fortún, Luis Manzano, Beatriz Montero-Errasquín, Lucienne Costa-Frossard, Jaime Masjuan, Luisa María Villar, for the COVID-HRC group

## Abstract

**INTRODUCTION:** Despite the increasing evidence of the benefit of corticosteroids for the treatment of moderate-severe Coronavirus disease 2019 (COVID-19) patients, no data are available about the potential role of high doses of steroids for these patients.

**METHODS:** All consecutive confirmed COVID-19 patients admitted to a single center were selected, including those treated with steroids and an acute respiratory distress syndrome (ARDS). Patients were allocated to the high doses (HD, ≥250mg/day of methylprednisolone) of corticosteroids or the standard doses (SD, ≤1.5mg/kg/day of methylprednisolone) at discretion of treating physician. The primary endpoint was the mortality between both cohorts and secondary endpoints were the risk of need for mechanical ventilation (MV) or death and the risk of developing a severe ARDS.

**RESULTS:** 573 patients were included: 428 (74.7%) men, with a median (IQR) age of 64 (54–73) years. In HD cohort, a worse baseline respiratory situation was observed and male sex, older age and comorbidities were significantly more common. After adjusting by baseline characteristics, HD were associated with a higher mortality than SD (adjusted-OR 2.46, 95% CI 1.58 – 3.83, p<0.001) and with an increased risk of needing MV or death (adjusted-OR 2.50, p=0.001). Conversely, the risk of developing a severe ARDS was similar between groups. Interaction analysis showed that HD increased mortality exclusively in elderly patients.

**CONCLUSION:** Our real-world experience advises against exceeding 1-1.5mg/kg/day of corticosteroids for severe COVID-19 with an ARDS, especially in older subjects. This reinforces the rationale of modulating rather than suppressing immune responses in these patients.

**SUMMARY:** In patients with severe COVID-19, high doses of corticosteroids are associated with a higher mortality and risk of need for mechanical ventilation or death compared to standard doses. This deleterious effect is mainly observed in the elderly.

## INTRODUCTION

Coronavirus disease 2019 (COVID-19) is a respiratory infection caused by the severe acute respiratory syndrome coronavirus 2 (SARS-CoV-2), a novel emergent virus that was first recognized in Wuhan, China, and has since rapidly spread around the world [1].

Although most patients present a mild-moderate disease, almost one-third of patients are at high risk of developing a more severe disease due to an acute respiratory distress syndrome (ARDS), that may lead to the need for mechanical ventilation (MV) and admission to an Intensive Care Unit, or even death [2]. The underlying mechanisms of severe COVID-19 are related to systemic inflammatory responses that can lead to lung injury and multisystem organ dysfunction [2, 3]. Based on this assumption, systemic anti-inflammatory drugs have been proposed as an alternative treatment tool to avoid the SARS-CoV-2-induced inflammatory state and to reduce mortality in these patients [3–6]. A first randomized controlled, clinical trial showed evidence that dexamethasone given at moderate doses for a short period of time reduced mortality vs. usual care in hospitalized COVID-19 patients [7]. Nevertheless, there is no evidence to clarify whether higher doses may improve or worsen the outcomes, and many uncertainties remain unclear in this regard. In this scenario, we aimed to evaluate the efficacy of high doses (HD) of corticosteroids in regard to standard doses (HD) in hospitalized patients with COVID-19 who developed an ARDS.

## METHODS

### Study design

This single-center, retrospective, observational study was performed at Hospital Universitario Ramón y Cajal (HRC) in Madrid, the region of Spain with the highest incidence of confirmed cases of COVID-19 in March and April 2020. All adult patients admitted to HRC with highly suspected SARS-CoV-2 infection from the beginning of the pandemics to April 15, 2020, were initially selected. Eligible patients included all hospitalized adults with a positive, laboratory-confirmed test for SARS-CoV-2 developing an acute respiratory distress syndrome (ARDS), treated with steroids, for whom a predefined minimum dataset was available. The minimum dose for inclusion was methylprednisolone-equivalent dosages of at least 0.5 mg/kg for 2 or more consecutive days. Patients without ARDS or treated during admission with remdesivir, a current anti-viral treatment that proved to be effective for SARS-CoV-2 infection [8], were excluded.

The study was approved by the institutional ethics board of HRC. The need for informed consent from individual patients was waived due to its retrospective design.

### Data collection

Trained physicians reviewed electronic medical records and extracted data for the period between admission to discharge, death or June 22, 2020, whichever occurred first. Demographical, clinical, radiological, and laboratory information were recorded, including comorbidities, respiratory variables, and details of treatments administered for COVID-19. Two different cohorts of patients were established, depending on the dosages of steroids administered for ARDS:

- High dose (HD) of corticosteroids: short-term pulse therapy of methylprednisolone-equivalent from 250 mg/day to 1000 mg/day during one or more consecutive days.
- Standard dose (SD) of corticosteroids: methylprednisolone-equivalent dosages ranging from and including 0.5 mg/kg/day to 1.5 mg/kg/day.

The decision of treatment with one dosage or another was exclusively at the discretion of treating medical team, as evidence about the use of corticosteroids on COVID-19 was very low. Whenever a patient was treated with both range of doses, the allocation to one group or another was established selecting the doses administered before the outcome variable occurred (e.g. a patient initially treated with SD developing the outcome who later received a HD was considered SD). Whenever both dosages were administered before the outcome, patients were classified as HD.

The primary endpoint was the mortality between HD and SD patients. Secondary endpoints included: 1) a combined variable of need for mechanical or non-invasive ventilation (MV) and death and 2) the development of severe acute respiratory distress syndrome (ARDS), according to the Berlin Definition [9].

### Statistical analysis

Continuous variables were reported as mean (standard deviation) or median (interquartile range) depending on the distribution of data and were evaluated with a two-sample t test or the Wilcoxon rank-sum test, as appropriate. Categorical variables were described using absolute and relative frequencies and analyzed with a χ^2^ test. We conducted both unadjusted and multivariable logistic regression models to investigate the effect of both dosages on primary and secondary endpoints. The multivariable model was adjusted by potential confounding factors identified at baseline (sex, age, age-adjusted Charlson Comorbidity Index –CCI- [10] and peripheral oxygen saturation/fraction of inspired oxygen –SpO2/FiO2-ratio). To avoid reverse causality associations between treatments and outcomes, we excluded for analysis those patients receiving the first dose of corticosteroids the same day or after the endpoint of interest. Besides, in order to increase specificity by considering a sufficient effect of corticosteroids, only patients exposed to treatment at least 3 days before the event of interest were considered for analyses. We also performed sensitivity analyses to validate the strength of findings. Results from all multivariable analyses are reported as odds ratios (ORs) with 95 % confidence intervals (CIs). We further analyzed interaction between both groups and age. All analyses were conducted using Stata^®^ 14 (StataCorp, College Station, TX, USA) and were two-tailed, with P <0.05 as the level of significance.

## RESULTS

### Patient characteristics

Eight hundred and thirty six consecutive hospitalized patients with a highly suspected SARS-CoV-2 infection were screened for inclusion. We first removed patients with a negative RT-PCR assay (n=48) or incomplete follow-up data (n=4). After excluding patients not receiving steroids (n=118), not developing ARDS (n=56) or those treated with remdesivir (n=37), 573 were finally included for analysis (Figure 1). By far the most chosen corticosteroid was methylprednisolone, administered to 568 (99.1%) patients. Three hundred and ninety six patients were in the SD cohort (69.1%), while 177 (30.9%) were treated with higher doses (HD). Further information about dosages, frequencies and duration of corticosteroids regimens is provided in Supplemental Material (Table 1). Median (IQR) time from onset of symptoms to admission, to ARDS and to first dose of steroids were 7 (4 – 9), 8 (6 – 11) and 10 (7 – 14) days, respectively, with no differences between both cohorts. Patients were followed for a median (IQR) of 21 (15 – 32) days. Baseline demographics, clinical data, radiological and laboratory findings are shown in Table 1. The median age of the study population was 64 years and HD patients were older compared to SD patients (67 vs. 64 years, respectively, p=0.03). There were significantly fewer men in the SD cohort (70.2%) than in the HD group (84.8%) (p<0.001). Relevant comorbidities were frequent in both groups. The age-adjusted CCI was mildly higher among HD patients (median [IQR] of 3 [2 – 5]) compared to SD patients (median [IQR] of 3 [1 – 4]) (p=0.009). In addition, patients treated with HD of steroids at the onset of ARDS presented a worse respiratory situation, with a lower SpO2/FiO2 ratio compared to SD patients (271 vs. 277 respectively, p=0.011). Baseline characteristics of patients treated with corticosteroids at least 3 days before MV (n=396) and severe ARDS (n=210), selected for analysis of secondary endpoints, are shown in Supplemental Material (Tables 2 and 3 of Supplemental Material).

**Table 1.** Baseline Demographics, Clinical Data and Radiological Findings.

|  |  |  |  |  |
| --- | --- | --- | --- | --- |
|  | **All patients**<br>**(n=573)** | **Standard dose**<br>**(n=396)** | **High dose**<br>**(n=177)** | **p Value** |
| Age (years), median (IQR) | 64 (54 – 73) | 64 (54 – 72) | 67 (55 – 76) | **0.03** |
| Men, No. (%) | 428 (74.7) | 278 (70.2) | 150 (84.8) | **<0.001** |
| Comorbidities, No. (%) |  |  |  |  |
| None | 126 (22) | 89 (22.5) | 37 (20.9) | 0.18 |
| Hypertension | 268 (46.8) | 179 (45.2) | 89 (50.3) | 0.26 |
| Diabetes | 113 (19.7) | 79 (20) | 34 (19.2) | 0.84 |
| Obesity<sup>a</sup> | 226 (39.4) | 162 (40.9) | 64 (36.2) | 0.28 |
| Cardiovascular disease<sup>b</sup> | 99 (17.3) | 63 (15.9) | 36 (20.3) | 0.20 |
| Chronic renal disease | 45 (7.9) | 28 (7.1) | 17 (9.6) | 0.30 |
| Chronic liver disease<sup>c</sup> | 36 (6.3) | 23 (5.8) | 13 (7.3) | 0.48 |
| Chronic lung disease<sup>d</sup> | 94 (16.4) | 62 (15.7) | 32 (18.1) | 0.47 |
| Autoimmune disease | 26 (4.5) | 19 (4.8) | 7 (4) | 0.54 |
| Active malignancy | 41 (7.2) | 23 (5.8) | 18 (10.2) | 0.06 |
| Malignancy in remission | 57 (10) | 35 (8.8) | 22 (12.4) | 0.19 |
| Organ transplant | 9 (1.6) | 5 (1.3) | 4 (2.3) | 0.38 |
| HIV infection | 5 (0.9) | 5 (1.3) | 0 | 0.13 |
| Dementia<sup>e</sup> | 25 (4.4) | 21 (5.3) | 4 (2.3) | 0.10 |
| CCI score, median (IQR) | 3 (1 – 4) | 3 (1 – 4) | 3 (2 – 5) | **0.009** |
| Time from symptoms onset to admission (days), median (IQR) | 7 (4 – 9) | 7 (4 – 9) | 7 (4 – 10) | 0.96 |
| Time from symptoms onset to ARDS (days), median (IQR) | 8 (6 – 11) | 8 (6 – 11) | 8 (6 – 12) | 0.53 |
| SpO<sub>2</sub>/FiO<sub>2</sub> ratio<sup>f</sup>, median (IQR) | 274 (188 – 375) | 277 (218 – 391) | 271 (180 – 332) | **0.011** |
| Pneumonia at admission |  |  |  |  |
| None | 15 (2.6) | 10 (2.5) | 5 (2.9) |  |
| Unilateral | 68 (11.9) | 49 (12.4) | 19 (10.7) | 0.84 |
| Bilateral | 490 (85.5) | 337 (85.1) | 153 (86.4) |  |
| Bacterial co-infection, No. (%) | 99 (17.3) | 65 (16.4) | 34 (19.2) | 0.41 |

Abbreviation: ARDS: acute respiratory distress syndrome; CCI: age-adjusted Charlson Comorbidity Index; IQR: interquartile range; No: number; SpO<sub>2</sub>/FiO<sub>2</sub>: pulse oximetry-based peripheral blood oxygen saturation/fraction of inspired oxygen.
<sup>a</sup>Defined as a body mass index $\geq 30$ kg/m<sup>2</sup>.
<sup>b</sup>Heart failure, myocardopathy, ischemic heart disease, and moderate-severe valvular heart disease.
<sup>c</sup>Chronic hepatitis, cirrhosis, hepatic steatosis, non-alcoholic steatohepatitis.
<sup>d</sup>Chronic obstructive pulmonary disease, obstructive sleep apnea-hypopnea syndrome, asthma, and diffuse interstitial lung disease.
<sup>e</sup>Includes developmental disabilities.
<sup>f</sup>Measured at development of ARDS.

**Table 2.** Primary and secondary outcomes among HD of corticosteroids during admission.

|  |  |  |  |  |  |  |  |
| --- | --- | --- | --- | --- | --- | --- | --- |
|  |  | Unadjusted |  |  | Multivariate model |  |  |
| Outcomes<sup>a</sup> | Number at risk<sup>b</sup> | OR | 95% CI | P value | OR | 95% CI | P value |
| Primary endpoint |  |  |  |  |  |  |  |
| **Death** | 573 | 2,79 | 1.87 – 4.15 | **<0.001** | 2.46 | 1.58 – 3.83 | **<0.001** |
| **Secondary endpoint** |  |  |  |  |  |  |  |
| **Need for MV or death** | 396 | 2.25 | 1.39 – 3.62 | **0.001** | 2.50 | 1.49 – 4.20 | **0.001** |
| **Severe ARDS** | 210 | 0.57 | 0.21 – 1.57 | 0.28 | 0.60 | 0.21 – 1.69 | 0.33 |

Abbreviation: ARDS: acute respiratory distress syndrome; MV: mechanical or non-invasive ventilation.
<sup>a</sup>Comparisons are performed with standard dose (SD) of steroids as reference.
<sup>b</sup>Only patients who received steroids at least 3 days before outcome were considered for analysis.

**Figure 1.**
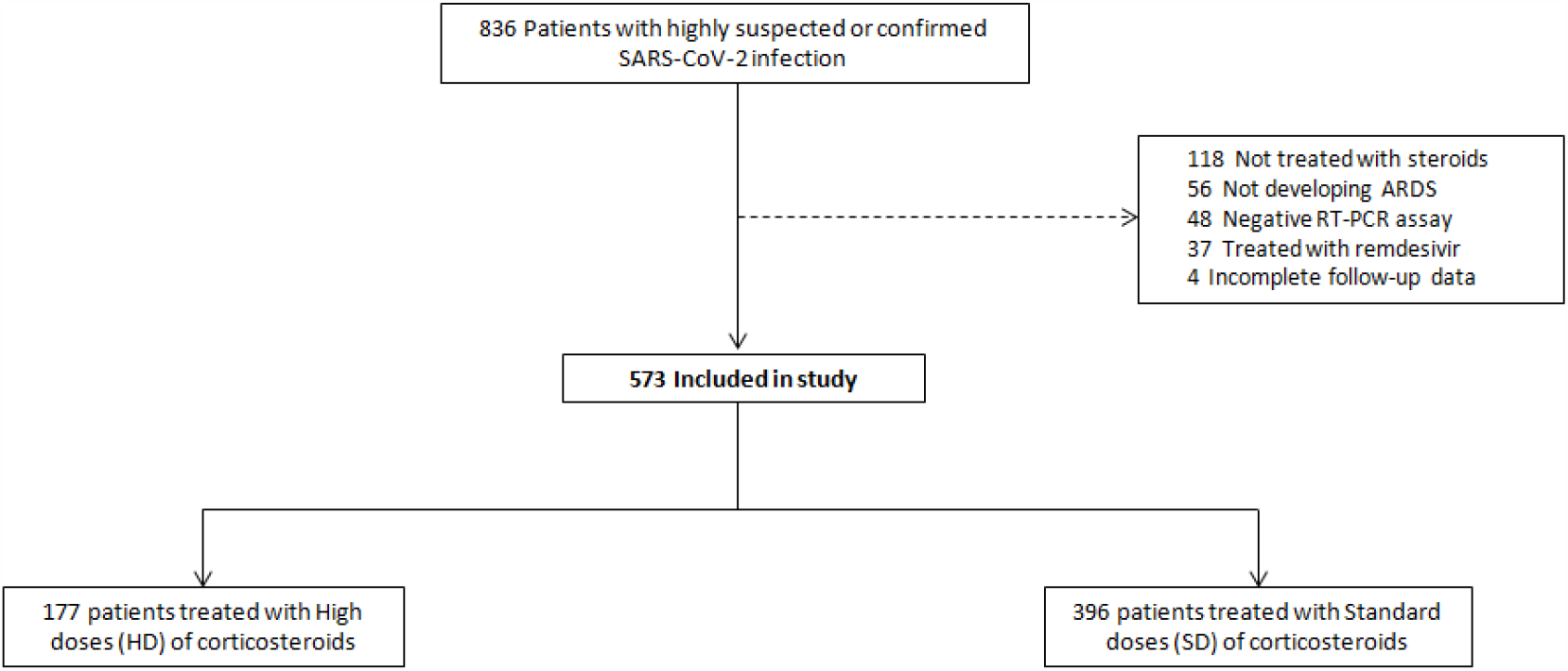
Patient inclusion and stratification. Abbreviations: ARDS, acute respiratory distress síndrome; RT-PCR, reverse transcriptase–polymerase chain reaction; SARS-CoV-2, severe acute respiratory syndrome coronavirus 2.

During admission, several treatments were administered to patients, being hydroxychloroquine the most frequent (98.1%), followed by lopinavir/ritonavir (92.5%) and antibiotics (89.2%), with no significant differences between both cohorts. Tocilizumab, a monoclonal antibody inhibiting interleukin-6 (IL-6) receptor, was equally given to both groups (40.9% among the SD cohort and 42.4% among the HD group). Only when considering rarely administered treatments, significant differences between HD and SD were observed (more anakinra and less interferon-β 1b among the HD group) (Table 4 of Supplemental Material).

### Outcomes

A final admission outcome (discharge or death) was recorded in 564 out of 573 (98.4%) patients. Overall mortality after a median (IQR) of 16 (9 – 26) days of admission was 24.8%, and was twice more frequently in the HD cohort than in the SD group (39% vs. 18.6%, respectively). The unadjusted logistic regression model showed a significantly higher risk of death for patients with an ARDS receiving HD of corticosteroids compared to patients treated with SD (OR 2.79, 95% CI 1.87 – 4.15, p<0.001). After adjusting by sex, age, CCI and SpO2/FiO2, results remained similar (adjusted OR 2.46, 95% CI 1.58 – 3.83, p<0.001). Among 396 patients treated at least 3 days before outcome, HD corticosteroids were associated with an increased risk of need for MV or death (adjusted-OR 2.50, 95% CI 1.49 – 4.20, p=0.001) compared to SD patients. No significant differences between both groups were observed in the risk of developing a severe ARDS (Table 2). We performed sensitivity analyses excluding patients hospitalized for no longer than seven days or those treated with less than five days of corticosteroids and all results remained practically unchanged.

As age is a strong prognostic factor, we further analyzed the effect of this variable in primary and secondary outcomes. Figure 2 shows the impact of HD of corticosteroids on mortality depending on the age of patients. The risk of death significantly increased in patients older than 65 years when compared to SD patients, being this effect attenuated in younger individuals. Age and HD of corticosteroids showed a trend towards a significant interaction between both (p=0.058). The interaction between age and HD in the risk of need for VM or death was not significant (Figure 1 of Supplemental Material).

**Figure 2.**
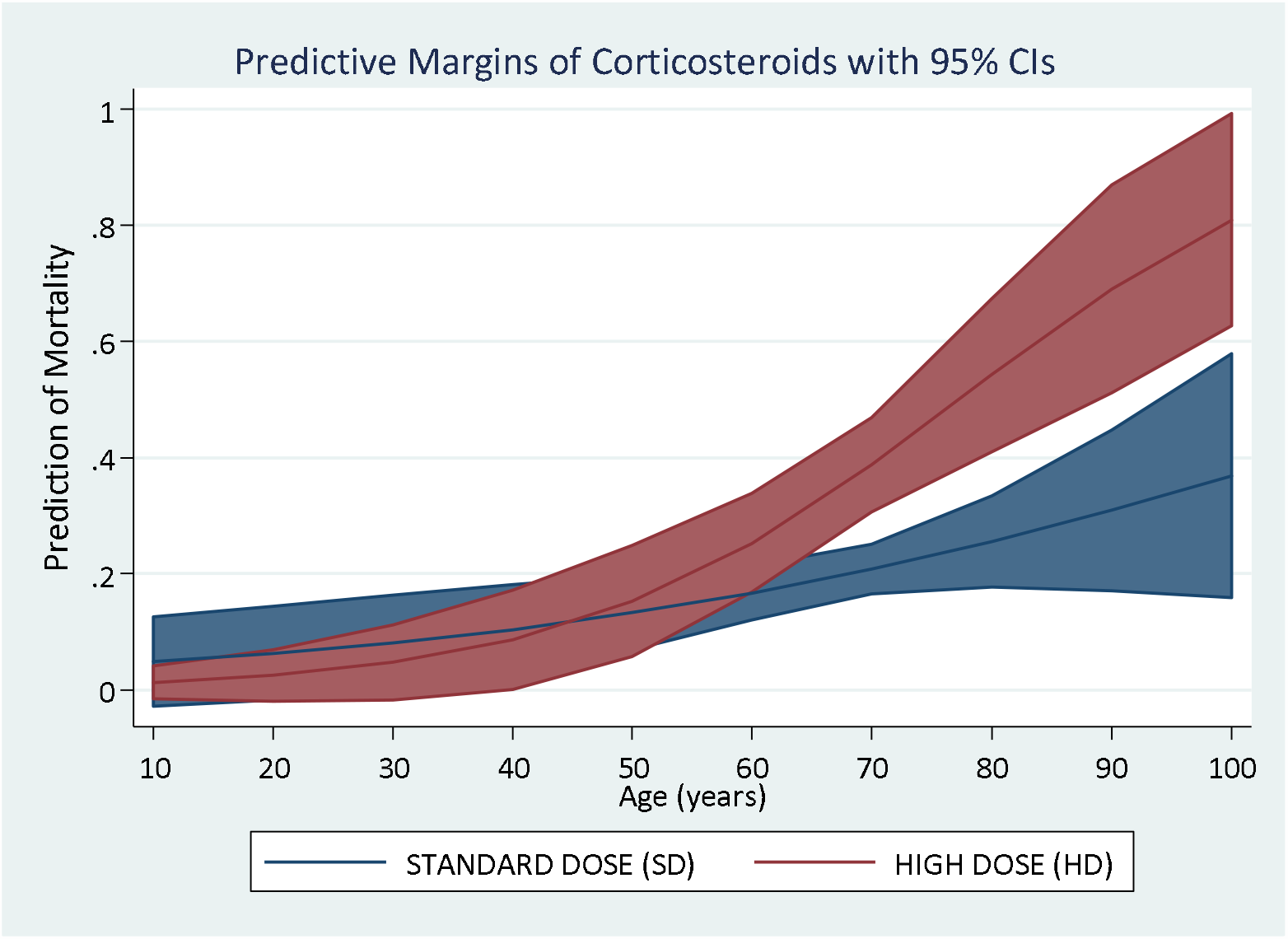
Interaction of age with both dosages of corticosteroids on mortality. The impact of high doses (HD) of corticosteroids is similar to standard doses (SD) in younger patients, but a a higher mortality is observed among the elderly. Abbreviations: CI, confidence interval.

## DISCUSSION

In this large observational study performed in Madrid, Spain, short-term high doses of corticosteroids, when compared with standard doses, were associated with an increased mortality need for MV or death in hospitalized patients with a SARS-CoV-2 infection developing an ARDS. This harmful effect was mainly observed in elderly patients, since young age attenuated the deleterious impact of higher doses.

Despite its known immunosuppressive effect, corticosteroids have been a treatment option for several bacterial, viral or even fungal infections, especially in the most severe, in order to regulate excessive immune responses causing tissue damage [11]. Steroids have been extensively used in severe cases of SARS-CoV-2 infection since the beginning of the pandemics [12]. The rationale was based on preliminary data suggesting that the immune response has two distinct phases [13]: the first triggered by the virus itself and characterized by mild constitutional and respiratory symptoms; the second consisting in an excessive inflammatory response leading to lung tissue injury and multisystem organ dysfunction in a minority of patients [14]. This hypothesis has been later supported by post-mortem case series, as histopathology findings reinforce the role of immune-mediated, rather than pathogen-mediated, pulmonary inflammation and death [15–16]. In this setting, therapeutic immunomodulation in severe COVID-19 was initially proposed on the basis of a strong scientific rationale [3–6, 17]. Several drugs aimed to limit immune-mediated injury in COVID-19 are consequently being investigated, including corticosteroids [18–21]. This treatment option is not new for coronavirus infections, as they were already used during the corresponding outbreaks of SARS and Middle East Respiratory Syndrome (MERS). However, results for SARS were either inconclusive or even harmful for patients [22] and no impact on mortality was observed for MERS. In fact, coronavirus RNA clearance was delayed [23]. Among hospitalized COVID-19 patients, steroids have been the anti-inflammatory treatment most evaluated [12]. Small cohort studies and case series yielded mixed results, reporting both positive [2, 24–26] and negative [27–29] outcomes. However, these works are prone to imbalances, indication bias and reverse causality. For example, a deleterious effect of steroids on influenza A-related critical illness was assumed until both baseline and time-dependent factors were considered in a later study [30]. For this reason, clinical trials are needed to provide the highest evidence for efficacy and safety. Preliminary results from a randomized, controlled, open-label trial comparing dexamethasone 6 mg given once daily for up to 10 days vs. usual care in hospitalized COVID-19 patients in the United Kingdom showed that patients who were allocated to receive dexamethasone had a reduced rate of mortality compared to those who concurrently allocated standard of care. This benefit was observed only in patients receiving invasive MV and those who required supplemental oxygen, but not in patients with mild infection [7].

For patients with an ARDS from any cause, general recommendations include the administration of corticosteroids at methylprednisolone-equivalent of 1mg/kg/day exclusively when a moderate or severe stage is observed [31]. Despite results from this initial clinical trial, there is uncertainty about whether higher doses of steroids in COVID-19 could provide a greater benefit, especially in the presence of a severe inflammation. To the best of our knowledge, no study has evaluated the effect of high dose pulse therapy (HD) compared to supportive care alone or standard doses (SD). Our results outline that HD are not associated with better outcomes, but with a higher risk of death and need for MV or death in comparison with SD. Although some differences in baseline characteristics were detected (male sex, older age and comorbidities were significantly more common among the HD cohort), appropriate adjustments were performed, concomitant treatments during admission were equally administered, and sensitivity analyses reinforced the robustness of our results.

These results emphasize that, even if some immunomodulatory or immunosuppressive treatments might be effective for severe COVID-19, the role of adaptive immunity is essential as viral dissemination is a key driver of severe disease. The optimal aim in severe COVID-19 might be to achieve a complicate balance between controlling the excessive innate immune hyperactivation, as well as recovering from the adaptive immune dysfunction, which has a critical role on clearing the viral infection and downregulating the innate immunity [3]. In fact, this may be the reason for the higher mortality observed in patients with moderate or severe immunosuppression when compared with general population [32–35], as well as the better outcomes whenever immunosuppression is non-severe [36]. A similar rationale may be applied to corticosteroids, aiming to modulate the immune hyperactivation without exerting a suppression of the adaptive immunity. In addition, higher doses could probably be associated with more and more serious adverse events, including concomitant infections [37], and clinical benefit should therefore exceed the increase of risks. Thus, standard doses given for the shorter time possible might be a better option for severe COVID-19 patients.

There are obvious limitations in a single-center study of this type. First, due to the observational character of the current work, potential confounding factors might have not been controlled and, therefore, conclusions must be taken with caution. In addition, the lack of randomization could have introduced indication bias, using HD of corticosteroids for more severe patients. Second, the range of doses used in the HD cohort contributed to some heterogeneity within this group. And finally, standardized care pathways and evidence-based treatment protocols for COVID-19 have not been established, and management might have been different between patients, introducing potential bias.

Despite these limitations, these results could have direct relevance to the evolving management of COVID-19 for treating physicians. In addition to the contribution made by the first clinical trial of dexamethasone at moderate doses [7], we add robust evidence to prevent from using high doses of corticosteroids for moderate or severe COVID-19 patients in order to avoid harmful effects.

In conclusion, among hospitalized patients with COVID-19 developing an ARDS, the administration of high doses of corticosteroids are associated with increased mortality and a higher risk of need for MV or death compared to standard doses. Thus, corticosteroids at 1 mg/kg/day of methylprednisolone-equivalent given for a short period might be more beneficial for these patients. Nevertheless, randomized, double-blind, controlled clinical trials are needed to provide stronger evidence in this regard.

## Data Availability

The datasets generated during and/or analysed during the current study are available from the corresponding author on reasonable request.

## CONFLICTS OF INTERES/FINANCIAL DISCLOSURES

The authors report no conflicts of interest.

## AUTHOR CONTRIBUTIONS

Drs Monreal, Sainz de la Maza and Villar had full access to all of the data in the study and take responsibility for the integrity of the data and the accuracy of the data analysis.

**Study concept and design:** Monreal, Sainz de la Maza, Muriel, Zamora, Villar.

**Acquisition, analysis or interpretation of data:** Monreal, Sainz de la Maza, Natera-Villalba, Beltrán-Corbellini, Rodríguez-Jorge, Fernández-Velasco, Walo-Delgado, Alonso-Canovas, Fortún, Manzano, Montero-Errasquín, Costa-Frossard, Masjuan, Villar.

**Drafting of the manuscript:** Monreal, Sainz de la Maza, Natera-Villalba, Villar.

**Critical revision of the manuscript for important intellectual content:** Monreal, Sainz de la Maza, Natera-Villalba, Beltrán-Corbellini, Rodríguez-Jorge, Fernández-Velasco, Walo-Delgado, Muriel, Zamora, Alonso-Canovas, Fortún, Manzano, Montero-Errasquín, Costa-Frossard, Masjuan, Villar.

**Statistical analyses:** Monreal, Muriel, Zamora.

**Administrative, technical or material support:** Monreal, Sainz de la Maza, Natera-Villalba, Beltrán-Corbellini, Rodríguez-Jorge, Fernández-Velasco, Walo-Delgado, Alonso-Canovas, Villar.

**Study supervision:** Villar.

**The COVID-HRC (COronaVIrus Disease 2019 – Hospital Ramón y Cajal) Group:** Masjuan, J; Fortún, J; Montero-Errasquín, B; Manzano, L; Máiz-Carro, L; Sánchez-García, EM; Hidalgo, F; Domínguez, AR; Pérez-Molina, JA; Sánchez-Sánchez, O; Comeche, B; Monge-Maillo, B; Barbero, E; Barbolla-Díaz, I; Aranzábal Orgaz, L; Cobo, J; Rayo, I; Fernández-Golfín, C; González, E; Rincón-Díaz, LM; Ron, R; Mateos-Muñoz, B; Navas, E; Moreno, J; Norman, J; Serrano, S; Quereda Rodríguez-Navarro, C; Vallés, A; Herrera, S; Mateos del Nozal, J; Moreno-Cobo, MA; Gioia, F; Concejo-Badorrey, MC; Ortiz Barraza, EY; Moreno, A; Chamorro, S; Casado, JL; Almonacid, C; Nieto, R; Diz, S; Moreno, E; Conde, M; Hermida, JM; López, M; Monreal, E; Sainz de la Maza, S; Costa-Frossard, L; Natera-Villalba, E; Chico-García, JL; Beltrán-Corbellini, Á; Rodríguez-Jorge, F; Fernández-Velasco, JI; Rodríguez de Santiago, E; Rita, CG; Iturrieta-Zuazo, I; De Andrés, A; Espiño, M; Vázquez, M; Fernández Lucas, M; Martínez-Sanz, J; García-Barragán, N; Buisán, J; Toledano, R; Alonso-Canovas, A; Pérez-Torre, P; Matute-Lozano, MC; López-Sendón, JL; García-Ribas, G; Corral, Í; Villar, LM.

## SUPPLEMENTARY MATERIAL

**Table S1.** Concomitant treatments during admission.

| **Table 1. Concomitant treatments during admission** |  |  |  |  |
| --- | --- | --- | --- | --- |
|  | **All patients**<br>**(n=573)** | **Standard dose**<br>**(n=396)** | **High dose**<br>**(n=177)** | **p Value** |
| Hydroxychloroquine, n (%) | 562 (98.1) | 386 (97.5) | 176 (99.4) | 0.11 |
| Lopinavir/ritonavir, n (%) | 530 (92.5) | 370 (93.4) | 160 (90.4) | 0.20 |
| Antibiotics, n (%) | 511 (89.2) | 347 (87.6) | 164 (92.7) | 0.07 |
| Azithromycin, n (%) | 457 (79.8) | 317 (80.1) | 140 (79.1) | 0.79 |
| Tocilizumab, n (%) | 237 (41.4) | 162 (40.9) | 75 (42.4) | 0.74 |
| Anakinra, n (%) | 8 (1.4) | 1 (0.3) | 7 (4) | **<0.001** |
| Interferon-β 1b, n (%) | 35 (6.1) | 32 (8.1) | 3 (1.7) | **0.003** |

**Table S2.**
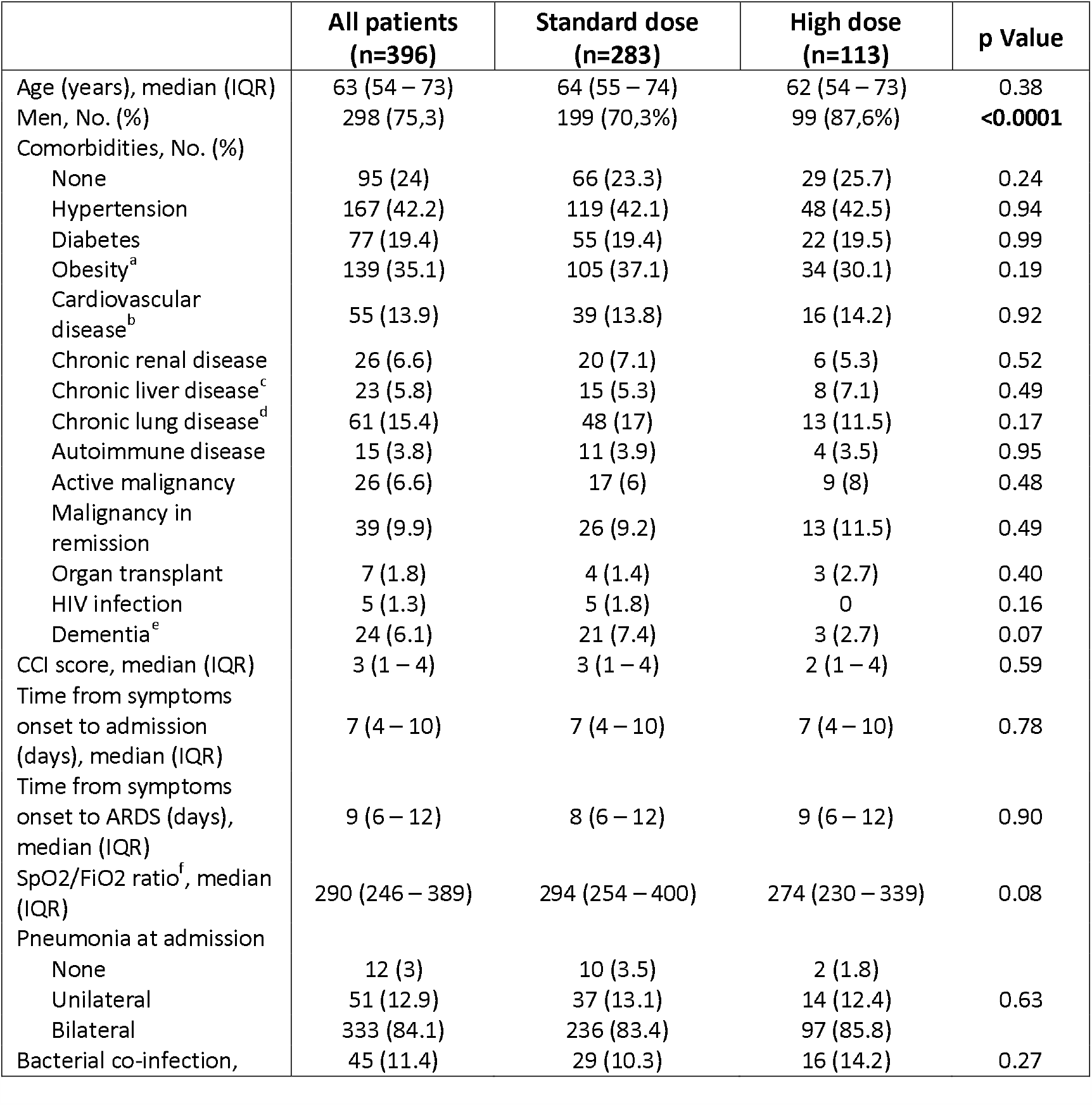
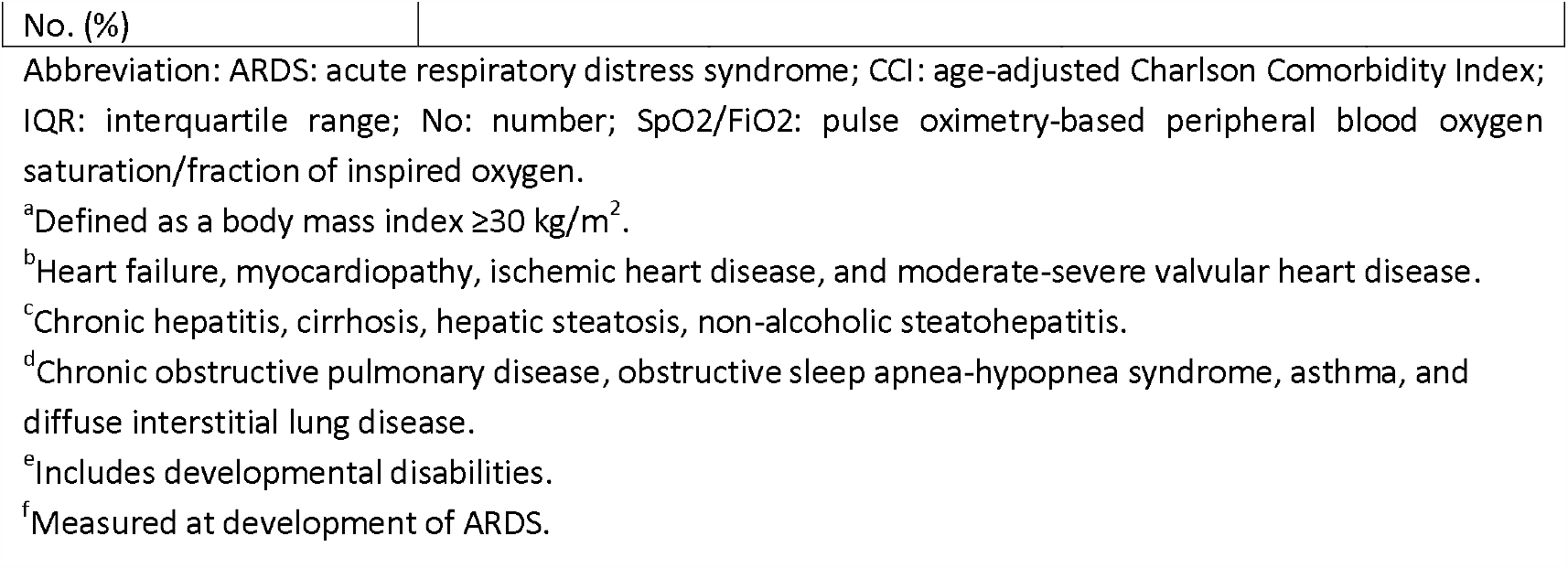
Baseline Demographics, Clinical Data and Radiological Findings in patients treated at least 3 days before MV.

| **Table 2. Baseline Demographics, Clinical Data and Radiological Findings in patients treated at least 3 days before MV** |  |  |  |  |
| --- | --- | --- | --- | --- |
|  | **All patients**<br>**(n=396)** | **Standard dose**<br>**(n=283)** | **High dose**<br>**(n=113)** | **p Value** |
| Age (years), median (IQR) | 63 (54 – 73) | 64 (55 – 74) | 62 (54 – 73) | 0.38 |
| Men, No. (%) | 298 (75.3) | 199 (70.3%) | 99 (87.6%) | **<0.0001** |
| Comorbidities, No. (%) |  |  |  |  |
| None | 95 (24) | 66 (23.3) | 29 (25.7) | 0.24 |
| Hypertension | 167 (42.2) | 119 (42.1) | 48 (42.5) | 0.94 |
| Diabetes | 77 (19.4) | 55 (19.4) | 22 (19.5) | 0.99 |
| Obesity<sup>a</sup> | 139 (35.1) | 105 (37.1) | 34 (30.1) | 0.19 |
| Cardiovascular disease<sup>b</sup> | 55 (13.9) | 39 (13.8) | 16 (14.2) | 0.92 |
| Chronic renal disease | 26 (6.6) | 20 (7.1) | 6 (5.3) | 0.52 |
| Chronic liver disease<sup>c</sup> | 23 (5.8) | 15 (5.3) | 8 (7.1) | 0.49 |
| Chronic lung disease<sup>d</sup> | 61 (15.4) | 48 (17) | 13 (11.5) | 0.17 |
| Autoimmune disease | 15 (3.8) | 11 (3.9) | 4 (3.5) | 0.95 |
| Active malignancy | 26 (6.6) | 17 (6) | 9 (8) | 0.48 |
| Malignancy in remission | 39 (9.9) | 26 (9.2) | 13 (11.5) | 0.49 |
| Organ transplant | 7 (1.8) | 4 (1.4) | 3 (2.7) | 0.40 |
| HIV infection | 5 (1.3) | 5 (1.8) | 0 | 0.16 |
| Dementia<sup>e</sup> | 24 (6.1) | 21 (7.4) | 3 (2.7) | 0.07 |
| CCI score, median (IQR) | 3 (1 – 4) | 3 (1 – 4) | 2 (1 – 4) | 0.59 |
| Time from symptoms onset to admission (days), median (IQR) | 7 (4 – 10) | 7 (4 – 10) | 7 (4 – 10) | 0.78 |
| Time from symptoms onset to ARDS (days), median (IQR) | 9 (6 – 12) | 8 (6 – 12) | 9 (6 – 12) | 0.90 |
| SpO<sub>2</sub>/FIO<sub>2</sub> ratio<sup>f</sup>, median (IQR) | 290 (246 – 389) | 294 (254 – 400) | 274 (230 – 339) | 0.08 |
| Pneumonia at admission |  |  |  |  |
| None | 12 (3) | 10 (3.5) | 2 (1.8) |  |
| Unilateral | 51 (12.9) | 37 (13.1) | 14 (12.4) | 0.63 |
| Bilateral | 333 (84.1) | 236 (83.4) | 97 (85.8) |  |
| Bacterial co-infection, | 45 (11.4) | 29 (10.3) | 16 (14.2) | 0.27 |

|  |  |
| --- | --- |
| No. (%) |  |
| Abbreviation: ARDS: acute respiratory distress syndrome; CCI: age-adjusted Charlson Comorbidity Index; IQR: interquartile range; No: number; SpO2/FiO2: pulse oximetry-based peripheral blood oxygen saturation/fraction of inspired oxygen. |  |
| <sup>a</sup>Defined as a body mass index $\geq 30$ kg/m<sup>2</sup>. | |
| <sup>b</sup>Heart failure, myocardopathy, ischemic heart disease, and moderate-severe valvular heart disease. |  |
| <sup>c</sup>Chronic hepatitis, cirrhosis, hepatic steatosis, non-alcoholic steatohepatitis. |  |
| <sup>d</sup>Chronic obstructive pulmonary disease, obstructive sleep apnea-hypopnea syndrome, asthma, and diffuse interstitial lung disease. |  |
| <sup>e</sup>Includes developmental disabilities. |  |
| <sup>f</sup>Measured at development of ARDS. |  |

**Table S3.** Characteristics of treatment with corticosteroids during admission.

| Table 3. Characteristics of treatment with corticosteroids during admission |  |  |  |
| --- | --- | --- | --- |
|  | Standard dose<br>(n=396) | High dose<br>(n=177) | P Value |
| Time from onset of symptoms to first dose (days), median (IQR) | 10 (7 – 14) | 11 (8 – 15) | 0.91 |
| Time with any corticosteroid (days), median (IQR) | 11 (7 – 16) | 11 (6 – 20) | 0.92 |
| Time with High Dose corticosteroids (days), median (range) | - | 3 (1 – 13) | - |
| Dosages per day (methylprednisolone-equivalent), n (%) |  |  |  |
| 250 mg | - | 69 (39) | - |
| 500 mg | - | 8 (4.5) | - |
| 1000 mg | - | 100 (56.6) | - |
| Patients treated with a second short-term course, n (%) | - | 8 (4.5) | - |

Abbreviation: IQR: interquartile range.

**Figure 1.**
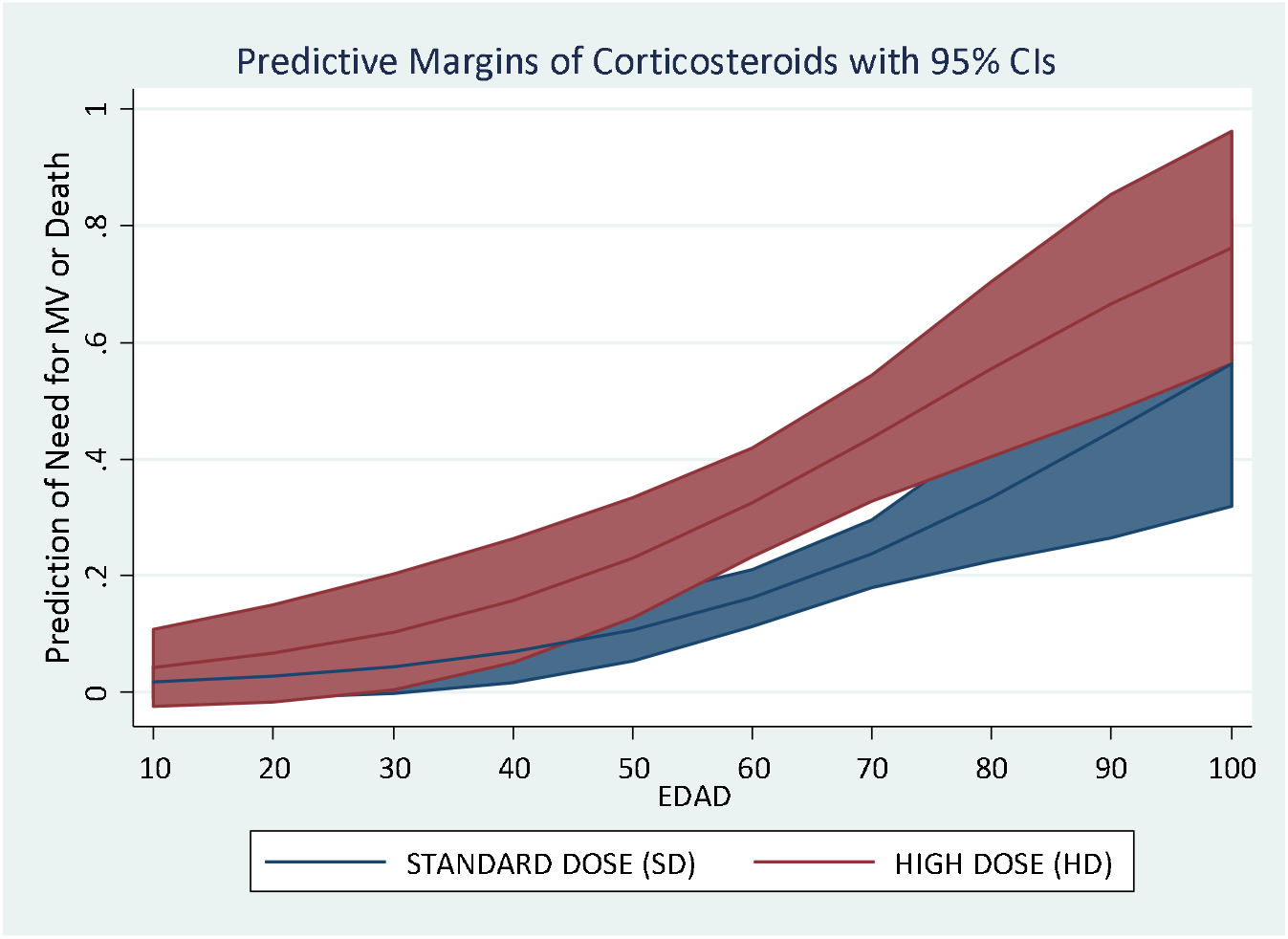

